# The Implementation, Diagnostic Yield and Clinical Outcome of Genetic Testing on an Inpatient Child and Adolescent Psychiatry Service

**DOI:** 10.1101/19004846

**Authors:** Aaron D. Besterman, Joshua Sadik, Michael J. Enenbach, Fabiola Quintero-Rivera, UCLA Clinical Genomics Center, Mark DeAntonio, Julian A. Martinez-Agosto

## Abstract

**Objective:** Diagnostic genetic testing is recommended for children with neurodevelopmental disorders (NDDs). However, many children with NDDs do not receive genetic testing. One approach to improve access to genetic services for these patients is to offer testing on the inpatient child and adolescent psychiatry (CAP) service.

**Methods:** We implemented systematic genetic testing on an inpatient CAP service by providing medical genetics education to CAP fellows. We compared the genetic testing rates pre- and post-education. We compared the diagnostic yield to previously published studies and the demographics of our cohort to inpatients who received genetic testing on other clinical services. We assessed rates of outpatient genetics follow-up post-discharge.

**Results:** The genetic testing rate on the inpatient CAP service was 1.6% (2/125) before the educational intervention and 10.7% (21/197) afterwards (OR = 0.13, 95% CI = 0.015- 0.58, *p* = 0.0015). Diagnostic yield for patients on the inpatient service was 4.3% (1/23), lower than previously reported. However, 34.8% (8/23) of patients had variants of unknown significance (VUSs). 39.1% (9/23) of children who received genetic testing while inpatients were underrepresented minorities, compared to 7.7% (1/13) of patients who received genetic testing on other clinical services (OR = 7.35, CI = 0.81-365.00, *p* = 0.057). 43.5% of patients were lost to outpatient genetics follow-up.

**Conclusion:** Medical genetics education for fellows on an inpatient CAP service can improve genetic testing rates. Genetic testing for inpatients may primarily identify VUSs instead of well-known NDD risk variants. Genetic testing on the inpatient CAP service may improve access to genetic services for underrepresented minorities, but assuring outpatient follow-up can be challenging.

## Introduction

Neurodevelopmental disorders (NDDs; referring to Intellectual Disability (ID), Autism Spectrum Disorder (ASD), and Global Developmental Delay (GDD) from here on) are genetically complex and heterogeneous^1,2^. Deletions or duplications of genomic regions >5 million base pairs (Mb) (e.g. copy number variants (CNVs)) that disrupt important neurobiological pathways are more common in individuals with NDDs than in the general population^3^. Similarly, gene-disrupting single base-pair changes called Single Nucleotide Variants (SNVs) are also more common in individuals with NDDs^3^. CNVs and SNVs that are associated with NDDs tend to be individually rare (<1% of the general population), *de novo* (e.g. not inherited from parents), and highly disruptive to key neurobiological pathways^4^. There are also so-called “syndromic” forms of NDDs that are associated with characteristic clinical features (e.g. specific dysmorphology or medical comorbidities) and are caused entirely by disruption of one gene^4^. For example, Fragile X (FX) syndrome is caused by disruption to the *FMR1* gene by a CGG trinucleotide repeat expansion of >200 copies.

Diagnostic genetic testing for patients with NDDs, using chromosomal microarrays (CMAs) to detect CNVs and polymerase chain reaction to detect FX syndrome, is now recommended by many professional medical organizations including the American College of Medical Genetics and Genomics (ACMG) and the American Academy of Child and Adolescent Psychiatry (AACAP)^5,6^. Diagnostic yield (e.g. the percent of patients with a positive genetic finding out of those who received genetic testing) using CMA is reported as 15-20%^7^. Whole Exome Sequencing (WES) to detect SNVs is often used as a second-line diagnostic test, but new guidelines are emerging suggesting its use as a first-tier test because of diagnostic yields of 30-50%^7^. There are many potential benefits to genetic testing and diagnosis, including improved reproductive counseling, enhanced community support from other affected families, improved prognostic information and appropriate medical monitoring, and the opportunity to participate in clinical research studies and trials^8,9^. There are also rare instances when a genetic diagnosis can have direct treatment implications^10–12^. The benefits of a genetic diagnosis will also grow as our knowledge about individual conditions expands. Despite these recommendations and potential benefits, genetic testing is often not offered to patients with NDDs by their physicians, including child and adolescent psychiatrists. The reason for this is multifactorial, but is likely due to a combination of insufficient training in genetics during residency and fellowship, limited awareness of the recommendations, limited awareness of potential benefits of genetic testing, limited time and a prioritization of other aspects of psychiatric care, barriers presented by insurance companies, and financial constraints ^13^.

The inpatient child and adolescent psychiatry (CAP) service is in many ways an ideal setting to provide genetic testing for patients with NDDs. Many patients with NDDs who are admitted to an inpatient psychiatric service have a multitude of psychiatric and medical comorbidities and lie at the severe end of the behavioral and functional impairment spectrum. A relatively high burden of pathogenic CNVs and SNVs have been observed in patients with NDDs and psychiatric comorbidities^14–16^, although the exact genetic architecture of these patients’ conditions in comparison to those without psychiatric comorbidities is not well-delineated.

Diagnostic genetic testing can also be integrated into the diagnostic work-up of patients on the inpatient service^6^. Therefore, in order to provide a much needed and underutilized diagnostic service that may improve outcomes for patients with NDDs, we attempted to implement systematic diagnostic genetic testing for patients with NDDs on our inpatient CAP service by educating CAP fellows in medical genetics. We then performed a retrospective analysis of the 12 months before our intervention with the 19 months after our intervention. Specifically, we analyzed genetic testing rates for patients with NDDs, diagnostic yield, and rates of outpatient follow-up compared to the 12-month period prior. We then compared our findings to previously published studies that reported diagnostic yield in patients with NDDs and comorbid psychiatric disorders ^14–16^.

## Methods

### Education Initiative

There are two inpatient CAP teams headed by separate attending physicians at our institution, the Resnick Neuropsychiatric Hospital at the University of California Los Angeles. One is run by first-year CAP fellows who rotate onto the service for an average of 4 months. The other is run by adult psychiatry residents completing their CAP training for an average of three weeks. Due to the high turnover rate of adult residents, the attending physician of this team (M.J.E.) played a primary role identifying patients with NDDs eligible for genetic testing. The formal education intervention was provided to the CAP fellows by A.D.B. It consisted of two parts: 1) All first-year CAP fellows were provided with an hour-long didactic session about the genetic architecture of NDDs and the rationale for genetic testing in patients with NDDs at the beginning of their academic year and 2)This was followed by a second, informal, didactic session during orientation for their inpatient CAP rotation, where they were refreshed on the benefits of genetic testing and introduced to the specific genetic testing protocol (available upon request) that we developed for the inpatient CAP service.

### Study Design and Protocol

Throughout the entire implementation process, the genetic psychiatry service (A.D.B) served as a consultant to the inpatient service for all questions regarding genetic testing and was available to assist with all parts of the testing process, from identification and evaluation of eligible patients to post-discharge follow-up. The genetic psychiatry service is a pilot clinical service at our institution with the goal of increasing the availability of genetics services to a wider range of psychiatry patients. We created a list of absolute and relative indications for genetic testing. Absolute indications were ID, GDD, ASD and childhood-onset schizophrenia (COS). Relative indications included epilepsy, severe psychopathology with congenital malformations, and high family loading of severe psychopathology suggesting possible mendelian inheritance. For the first 12 months after the education intervention, only FX testing and Single Nucleotide Polymorphism (SNP) CMA (patient/proband only) were recommended for all patients. From 12 months to 19 months post-intervention, we offered reflex trio (patient and both parents) WES when CMA and FX were negative or detected a variant of unknown significance (VUS) based on evidence of improved diagnostic yield^7^. Signed informed consent for CMA and WES was obtained from parents or guardians. Risks associated with genetic testing were discussed with families, including the risk of finding unrelated but actionable genetic variants per ACMG guidelines^17^, misattributed parentage, consanguinity, and potential impact on life and disability insurance eligibility. The limitations of genetic testing were also discussed. Orders for the genetic testing were placed by the fellow or resident who were assigned to the patient. Blood was drawn from the patients by UCLA Medical Center inpatient phlebotomists. Parents had their blood drawn at a UCLA-affiliated outpatient phlebotomy laboratory. Genetic analysis and interpretation were completed by the CLIA-certified UCLA Molecular Diagnostics Laboratory, as previously reported for WES^18^, with the addition of using the Genome Aggregation Database for variant analysis^19^. SNP CMA Diagnostic criteria at UCLA is the following: A CNV is reported when at least 25 adjacent oligonucleotide probes within a 50-200 kb segment or at least 50 adjacent oligonucleotide probes within a ≥200 Kb segment show a copy number change. The interpretation follows the ACMG guidelines^20^ for pathogenic variants, VUS (likely pathogenic), and VUS (no subclassification). VUS (likely benign) and benign variants are not reported. The SNP component of this microarray is designed to screen for regions of homozygosity (ROH) > 5 Mb. ROH can be indicative of uniparental disomy if within a chromosome, or an increased risk of a recessive disorder if identified across multiple chromosomes. WES results were discussed at UCLA Genomics Boards with A.D.B. and J.A.M.A. in attendance. SNV classification is determined by the Genomics Board based off of guidelines set by the ACMG and the Association for Molecular Pathology^21^. Outpatient follow-up appointments with medical genetics, genetic psychiatry, or genetic counseling were scheduled for each patient depending on specific patient factors. When genetic test results were finalized prior to patient discharge, they were returned to the family by the patient’s fellow or resident in consultation with either genetic psychiatry or a genetic counselor.

### Data Extraction and Chart Review

We obtained UCLA Institutional Review Board approval for chart review and data extraction on November 20, 2018 (IRB#16-001945). With the assistance of the UCLA Clinical Translational Sciences Institute Informatics Program, we extracted demographic information, medical, family and psychiatric history, medical and psychiatric diagnoses, genetic test results, and outpatient medical genetics encounters for patients admitted to our inpatient child and adolescent psychiatry service from July 1, 2016 – January 1, 2019 who carried the following diagnoses with the associated ICD-10 codes: Autism Spectrum Disorder (F84.0, F84.1, F84.2, F84.3, F84.4, F84.5, F84.8, F84.9), Intellectual Disability (F70, F71, F72, F73, F78, F79), Unspecified disorder of psychological development (F89), Childhood onset schizophrenia (F20) and/or GDD (F88). Information was extracted from the UCLA CareConnect (EPIC) Electronic Health Record (EHR) System using the Integrated Clinical and Research Data Repository (xDR). xDR is a clinical data warehouse system containing data from the UCLA EHR linked with older legacy systems and other sources. The requested data was extracted with xDR using SQL and output into encrypted comma-separated values files. All data was manually reviewed and curated by A.D.B. and J.S.

### Data Analysis

Fisher’s exact test was used to compare rates of genetic testing in the pre-intervention 12 month period to rates of genetic testing in the post-intervention 19 month period. Fisher’s exact test was also used to compare rates of underrepresented minorities in those patients who received genetic testing while on the inpatient unit to those patients who received genetic testing from other clinical services. Fisher’s exact test was used instead of the chi-squared test because there were less than 5 individuals in the samples^22^. All analyses were performed using R Studio (RStudio: Integrated development environment for R, Version 1.2.1335, Boston, MA).

## Results

### Rates of Genetic Testing and Diagnostic Yield

A total of 125 patients with eligible diagnoses were admitted to the inpatient service in the pre-intervention year. Of those patients, only two (1.6%) received genetic testing while on the inpatient service. One had both a pathogenic or likely pathogenic (P/LP) variant and a VUS, while the other had no findings (Table 1, Table S1, available online). A total of 197 patients with eligible diagnoses were admitted to the inpatient service in the post-intervention 19-month period. Twenty-one patients (10.7%) received genetic testing. The increase in genetic testing rate between the pre- and post-intervention years was statistically significant (OR = 7.26, 95% CI = 1.72-65.12, *p* = 0.0015). In the combined 31 months that included the pre- and post-intervention periods, 23 patients received genetic testing while on the inpatient service with a diagnostic yield for P/LP variants of 4.3% (Tables 1, S1, S2). 34.8% of tested patients had at least one VUS. Inheritance status of VUSs found on CMA are unknown because parental testing was not completed. We additionally assessed the number of eligible patients who were admitted to the inpatient unit who had received genetic testing in clinical settings at UCLA (e.g. outpatient neurology or medical genetics clinics) outside of their admission. 13 out of 322 patients (4.0%) with an eligible diagnosis who had been admitted to the inpatient service received genetic testing in clinical settings outside of the inpatient service (Table 2). Testing approaches in these settings were inconsistent, not always in accordance with current recommendations (e.g. did not provide FX and CMA testing to all patients), or was targeted based on clinical presentation (e.g. *PTEN* sequencing for patients with ASD and macrocephaly). Klinefelter’s syndrome was identified in one patient and VUSs were found in two patients. We assessed demographic distribution between the patients who received testing while inpatient compared to those who received testing from another clinical service. Only 7.7% of patients tested on other clinical services were from underrepresented minorities, while 39.1% of patients tested while on the inpatient service were underrepresented minorities (OR = 7.35, CI = 0.81-365.00, *p* = 0.057)

**Table 1:** Demographics, Genotype, Phenotype and Clinical Care of Inpatients who Received Genetic Testing While on the Inpatient Service

| Patient | Age/Sex/Ethnicity | Reason for Hospitalization | NDD<br>(Historical) | Psychiatric<br>Comorbidities | Medical Comorbidities | Genetic<br>Tests | Test Results<br>(Classification) | Ordering<br>Clinician | Genetics<br>Follow-up |
| --- | --- | --- | --- | --- | --- | --- | --- | --- | --- |
| 12 Months Prior to Education Intervention |  |  |  |  |  |  |  |  |  |
| 1 | 10/M/H | Assaultive and sexually aggressive behavior with frequent tantrums | ID | TS, OCD, ADHD, ODD | Situs inversus | CMA, FX | 12p12.1 del (LP); 15q26.2, 15q26.2q26.3 and 15q26.3 duplication (VUS) | CAPF | LTF |
| 2 | 17/F/H | Anxiety, depression, SI | ID, ASD | UAD, UMD, conversion disorder | Congenital nystagmus, eosinophilic gastritis | CMA | Negative | CAPF | LTF |
| 19 Months After Education Intervention |  |  |  |  |  |  |  |  |  |
| 3 | 16/M/C | Aggression | ID, ASD, (GDD) | IED | Dysmorphic facies | CMA, FX | Negative | APR | MG, WES Negative |
| 4 | 15/M/H | Court-ordered evaluation for aggressive and erratic behavior | ASD | ADHD, CD | Clubbed feet, epilepsy, PNES, high pain tolerance, dysmorphic facies | CMA, FX | Negative | CAPF | LTF |
| 5 | 16/M/C | Assaultive behavior at outpatient clinic | ASD | OCD, IED | Constipation | CMA, FX | Negative | APR | MG, WES denied |
| 6 | 17/M/C | Acute decline in functioning, aggression, auditory and visual hallucination, SI, HI | ID (GDD) | UPD, OCD, ADHD | History of brain cyst and meningitis | CMA, FX | 15q11.2 dup, BP1- BP2 (VUS); 11q24.3q25 dup (VUS) | CAPF | MG, parental CMA denied |
| 7 | 17/M/C | Aggression, SIB | ASD | ADHD, BD, UAD, IED |  | CMA, FX | Negative | CAPF | Not indicated<sup>a</sup> |
| 8 | 17/M/C | Rapid behavioral decompensation including increased OCD behaviors and gross thought disorganization | ASD | MDD, OCD, TS, UAD | Epilepsy, Burkitt's lymphoma, h/o DVT | CMA, FX | Negative | CAPF | LTF |
| 9 | 11/M/H | Aggression | ID, ASD, (GDD) | IED, UMD | Tardive dyskinesia | CMA, FX | 5q14.3 deletion (VUS) | APR | LTF |
| 10 | 15/M/C | Aggression, repeated elopement, SI | ID, ASD (GDD) | UPD | FASD, Brain injury of prematurity, pineal cyst, bowel obstruction, absence seizures | CMA, FX | Negative | APR | LTF |
| 11 | 13/F/C | SI and SIB | ASD | ODD, ADHD, UMD | FASD | CMA, FX | Negative | APR | LTF |
| 12 | 12/M/C | Aggression, impulsivity, and SIB | ASD, (GDD) | IED | Tardive dyskinesia | CMA, FX | Negative | CAPF | LTF |
| 13 | 15/M/C | Aggression and SIB | ASD, (GDD) | IED | GERD, eosinophilic esophagitis, severe constipation, rectal prolapse | CMA, FX | Negative | CAPF | Referred to outside MG |
| 14 | 15/M/AS | Psychosis and aggression | COS | GAD | Eczema | CMA, FX, WES | SHANK2 missense (VUS) | CAPF | Not indicated<sup>a</sup> |
| 15 | 16/M/C | Aggression and SIB | ID, ASD | - | Epilepsy | CMA | Negative | CAPF | Not indicated<sup>a</sup> |
| 16 | 16/F/C | Mutism with decline in ADLs, erratic behavior, possible catatonia | ID | UPD, ADHD, catatonia | - | CMA | 8p23.1p22 deletion (VUS) | CAPF | MG, annual f/u |
| 17 | 16/F/C | Decline in ADLs, SI with erratic and hypersexual behavior | - | UPD, MDD, UAD | Marfanoid habitus, dysmorphic facies | CMA, FX, WES | Negative | CAPF | Genetic psychiatry, enrolled in WGS study |
| 18 | 11/M/AA | Grossly psychotic behavior | COS | - | Atopic dermatitis | CMA, WES | Negative | CAPF | Referred to outside MG |
| 19 | 15/F/C | Aggression, functional decline | ASD | UPD, ADHD, OCD | - | CMA, FX, WES | SLC6A17 missense (VUS) | CAPF | Genetic psychiatry |
| 20 | 16/M/H | Aggression | ID, ASD | IED | Infantile spasms, focal cortical dysplasia | CMA, FX | 5q35.1 duplication (VUS) | CAPF | LTF |
| 21 | 9/M/H | Aggression | ASD | IED | Constipation, urinary retention | CMA, FX, WES | Negative | APR | Genetic psychiatry |
| 22 | 6/M/C | Aggression, SIB | ASD | ADHD | Leukocytosis, UTI, hives, pectus excavatum | CMA | Negative | APR | MG |
| 23 | 10/M/AS | Aggression | ASD | IED | - | CMA | 5q33.3 duplication (VUS) | CAPF | LTF |

<sup>a</sup> = Genetic test results available prior to discharge; AA = African American; AS = Asian; ADHD = Attention Deficit Hyperactivity Disorder; ADLs = Activities of Daily Living; APR = Adult Psychiatry Resident; ASD = Autism Spectrum Disorder; BD = Bipolar Disorder; BP = Breakpoint; C = Caucasian; CMA = Chromosomal Microarray; CAPF = Child and Adolescent Psychiatry Fellow; CD = Conduct Disorder; COS = Childhood-Onset Schizophrenia; F = Female; FX = Fragile X Testing; GERD = Gastroesophageal Reflux Disease; GDD = Global Developmental Delay ID = Intellectual Disability; IED = Intermittent Explosive Disorder; GAD = Generalized Anxiety Disorder; H = Hispanic; HI = Homicidal Ideation; LP = Likely Pathogenic; M = Male; MG = Medical Genetics ; ODD = Oppositional Defiant Disorder; PNES = Psychogenic Nonepileptic Seizure; SI = Suicidal Ideation; SIB = Self-Injurious Behavior; TS = Tourette's Syndrome; UAD = Unspecified Anxiety Disorder; UMD = Unspecified Mood Disorder; UPD = unspecified psychotic disorder; UTI = Urinary Tract Infection; VUS = Variant of Unknown Significance; WES = Whole Exome Sequencing; WGS = Whole Genome Sequencing

**Table 2:** Demographics, Genotype, and Phenotype of Inpatients who Received Genetic Testing in Other Clinical Settings

| Patient | Age/Sex/Ethnicity | Reason for Hospitalization | NDD (Historical) | Psychiatric Comorbidities | Medical Comorbidities | Genetic Tests | Test Results (Classification) | Ordering Service |
| --- | --- | --- | --- | --- | --- | --- | --- | --- |
| 24 | 7/M/C | Aggression | ASD | ADHD | - | FX, CMA | 15q11.2 dup, BP1-BP2 (VUS) | OP neurology |
| 25 | 7/M/C | Aggression | ASD | ADHD | Epilepsy, tracheomalacia | CMA | Negative | OP neurology |
| 26 | 12/M/C | Aggression, Impulsivity, HI | ASD | MDD, ADHD, IED, unspecified learning disorder | - | CMA, WES | Negative | OP MG |
| 27 | 14/M/C | Tantrums | ASD | IED | Normocytic anemia, gingivitis | Karyotype | Negative | OP endocrinology |
| 28 | 10/M/C | Aggression, SIB | ID, ASD, (GDD) | PANS, IED, ADHD, UAD | - | FX, CMA, WES | Negative | OP MG |
| 29 | 14/M/C | Aggression | ID, ASD, (GDD) | - | Constipation | MTHFR | Negative | Adolescent medicine |
| 30 | 16/F/C | Bizarre behavior and agitation | - | Schizophrenia, GAD, social phobia | Chronic pancreatitis, hyperlipidemia | CMA | Negative | IP neurology |
| 31 | 14/M//C | Irritability and head-banging | ID, ASD | IED, pica | Epilepsy, Septo-optic dysplasia | FX, CMA | Negative | IP neurology |
| 32 | 15/M/C | Depression and emotional dysregulation | ASD | GAD, UDD, ADHD | Obesity, OSA, Migraines | PTEN | Negative | OP MG |
| 33 | 17/F/H | Suicide attempt | ASD | ADHD, PDD | Obesity, PCOS, OSA, headaches | FX, PTEN | Negative | OP neurology |
| 34 | 17/M/C | Aggression | ID, ASD, (GDD) | OCD | Speech apraxia | FX, CMA, WES | AFF3 (VUS) | OP MG |
| 35 | 13/M/C | Suicidal ideation | ASD | ADHD, UDD | - | Karyotype, FX, CMA, WES, FDMA | 2q21 del (VUS)<br>2q21.1 dup (VUS)<br>8p22 triplication (VUS) | OP MG |
| 36 | 15/M/C | Depression, suicidal ideation, aggression | ASD | MDD, ADHD | Klinefelter Syndrome, GERD | Unknown | XXY (P) | Unknown |

ADHD = Attention Deficit Hyperactivity Disorder; ASD = Autism Spectrum Disorder; BP = breakpoint; C = Caucasian; CMA = Chromosomal Microarray; Del = deletion; Dup = duplication; FDMA = Familial Dysautonomia Mutational Analysis; FX = Fragile X Testing; GAD = Generalized Anxiety Disorder; GDD = Global Developmental Delay; GERD = Gastroesophageal Reflux Disease; H = Hispanic; HI = Homicidal Ideation; ID = Intellectual Disability; IED = Intermittent Explosive Disorder; IP = Inpatient; MG = Medical Genetics; MTHFR = Methylenetetrahydrofolate Reductase mutational testing; OP = Outpatient; OCD = Obsessive Compulsive Disorder; OSA = Obstructive Sleep Apnea; P = pathogenic PDD = Persistent Depressive Disorder; PANS = Pediatric Acute-onset Neuropsychiatric Syndrome; PCOS = Polycystic ovary syndrome; PTEN = Phosphatase and Tensin Homolog mutational testing; SIB = Self-Injurious Behavior; UAD = Unspecified Anxiety Disorder; UDD = Unspecified Depressive Disorder; VUS = Variant of Unknown Significance; WES = Whole Exome Sequencing

### Demographics and Clinical Outcomes

Eighteen out of 23 patients (78.3%) who received genetic testing on the inpatient service were male and 14/23 (60.9%) were non-Hispanic Caucasian (age range 6-17 years old). For the patients who received genetic testing from other clinical services, 11/13 (84.6%) were male and 12/13 (92.3%) were non-Hispanic Caucasian (age range 7-17 years old). Ethnicity was determined based on report in EHR. Most patients had multiple medical and psychiatric comorbidities (Table 1, Figure 1), were on multiple psychotropic medications (Table S3, available online), and had a significant family history of psychiatric illness (Table S4, available online). Attempts were made to coordinate outpatient genetics follow-up for all patients who received genetic testing, but 10/23 patients (43.5%) were still lost to outpatient follow-up (Figure 2). There were no instances where genetic testing altered medical management of the patients while on the inpatient service, but one patient was referred to a research study for whole genome sequencing.

**Figure 1:**
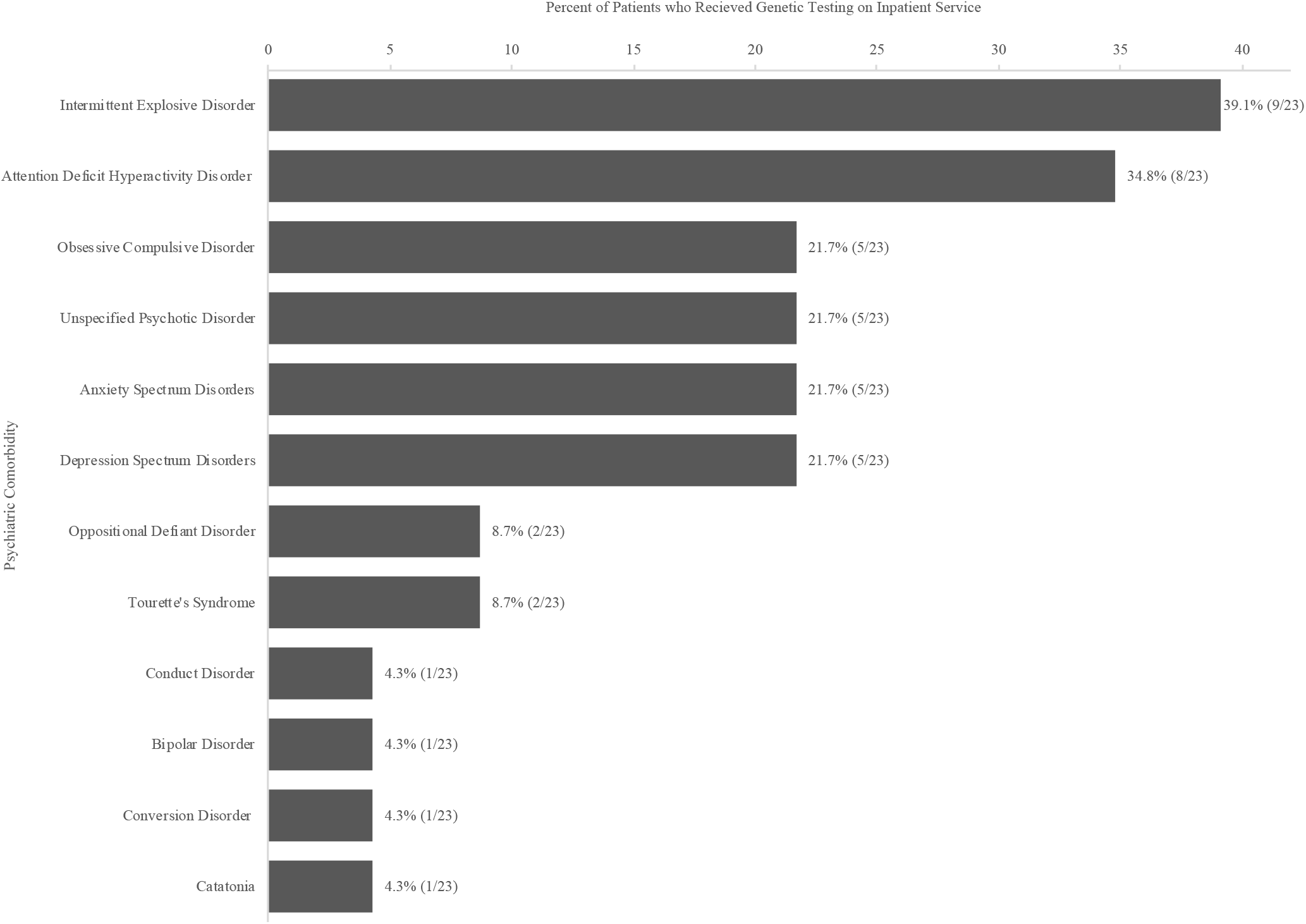
Psychiatric Comorbidities of Patients who Received Genetic Testing While on the Inpatient Child and Adolescent Psychiatry Service

**Figure 2:**
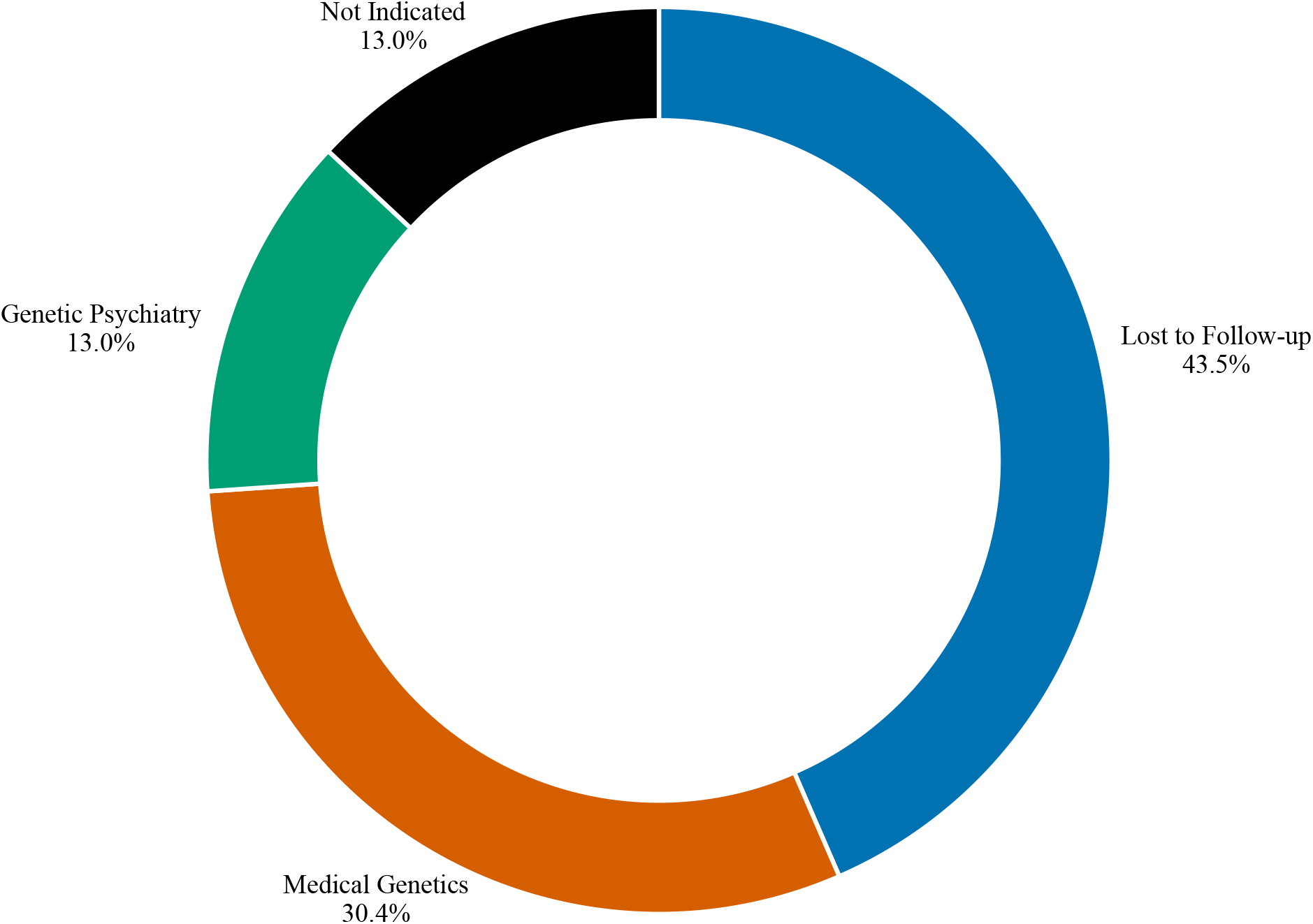
Outpatient Genetics Follow-up for Patients who Received Genetic Testing While on the Inpatient Service

## Discussion

### Genetic Test Results and Diagnostic Yield

When we designed the initial genetic testing protocol, we decided to expand eligibility criteria beyond current guidelines (e.g. ID, ASD, GDD) to include COS, complex psychiatric presentation and a family history enriched for psychiatric disorders. COS is a rare and severe NDD with a reported enrichment of known NDD-associated pathogenic CNVs compared to both matched, related controls and patients with adult-onset schizophrenia^23^, as well as disease-associated SNVs^24–26^. We were careful to include only those patients who had a well-documented history of psychosis-onset prior to the beginning of puberty. In one patient with COS, we identified a heterozygous missense VUS in *SHANK2* (Tables 1, Table S2, available online), which codes for a post-synaptic density protein highly expressed in glutamatergic synapses^27^. *SHANK2* missense variants have been linked to schizophrenia in case-control and family studies^27,28^, but was not identified as a schizophrenia risk gene in a recent, large trio WES study^29^. The father was unavailable for testing, so we were unable to conclude if it was *de novo* or paternally inherited, making it more difficult to conclude pathogenicity. Future reanalysis of these results (when more is known about the association of *SHANK2* to psychopathology) will likely be informative.

We also included patients who had a common psychiatric presentation without an NDD but with atypical comorbid features suggestive of a specific genetic syndrome. For example, one patient presented with adolescent-onset psychosis, depression and anxiety as well as a strikingly marfanoid habitus and dysmorphic facial features, suggestive of the 3q27.3 microdeletion syndrome^30^. However, the patient did not carry this deletion nor a pathogenic, exonic SNV in any genes within the region. We referred the patient to a whole genome sequencing study to try to identify a novel pathogenic variant. While we did not have any positive findings in the small number of patients included under these additional criteria, it is especially important that psychiatrists recognize the potential benefit of genetic testing in these populations because they are often the patient’s primary physician. In contrast, for patients with ASD, ID, or GDD, other specialists (e.g. medical geneticists) are commonly involved in clinical management.

The overall diagnostic yield for the 23 patients identified through our inpatient genetic testing protocol was 4.3%, based on a likely pathogenic deletion in 12p12.1 (Lamb-Shaffer Syndrome; MIM 604975) in patient 1 (Table 1). It is unknown if this variant is *de novo* as parents were unavailable for testing. To our knowledge, this is the first report of a patient with Lamb-Shaffer Syndrome with *situs inversus*, Tourette’s Syndrome, ADHD and OCD, although given the presence of an additional VUS that the patient carried, it is difficult to fully attribute the full phenotype to the 12p12.1 deletion ^31^. No P/LP variants were identified through WES or FX testing (Table 1). The low yield on FX testing is consistent with other recent studies in NDDs and supports the argument that FX testing may not be an appropriate first-tier test for all patients with NDDs ^32^, including on the inpatient service. By comparison, Viñas-Jones *et al*. successfully diagnosed FX Syndrome in five patients with NDDs and psychiatric co-morbidities by only testing those patients with clinical features consistent with FX Syndrome^15^ (Table 3), which may be a superior approach^32^.

**Table 3:**
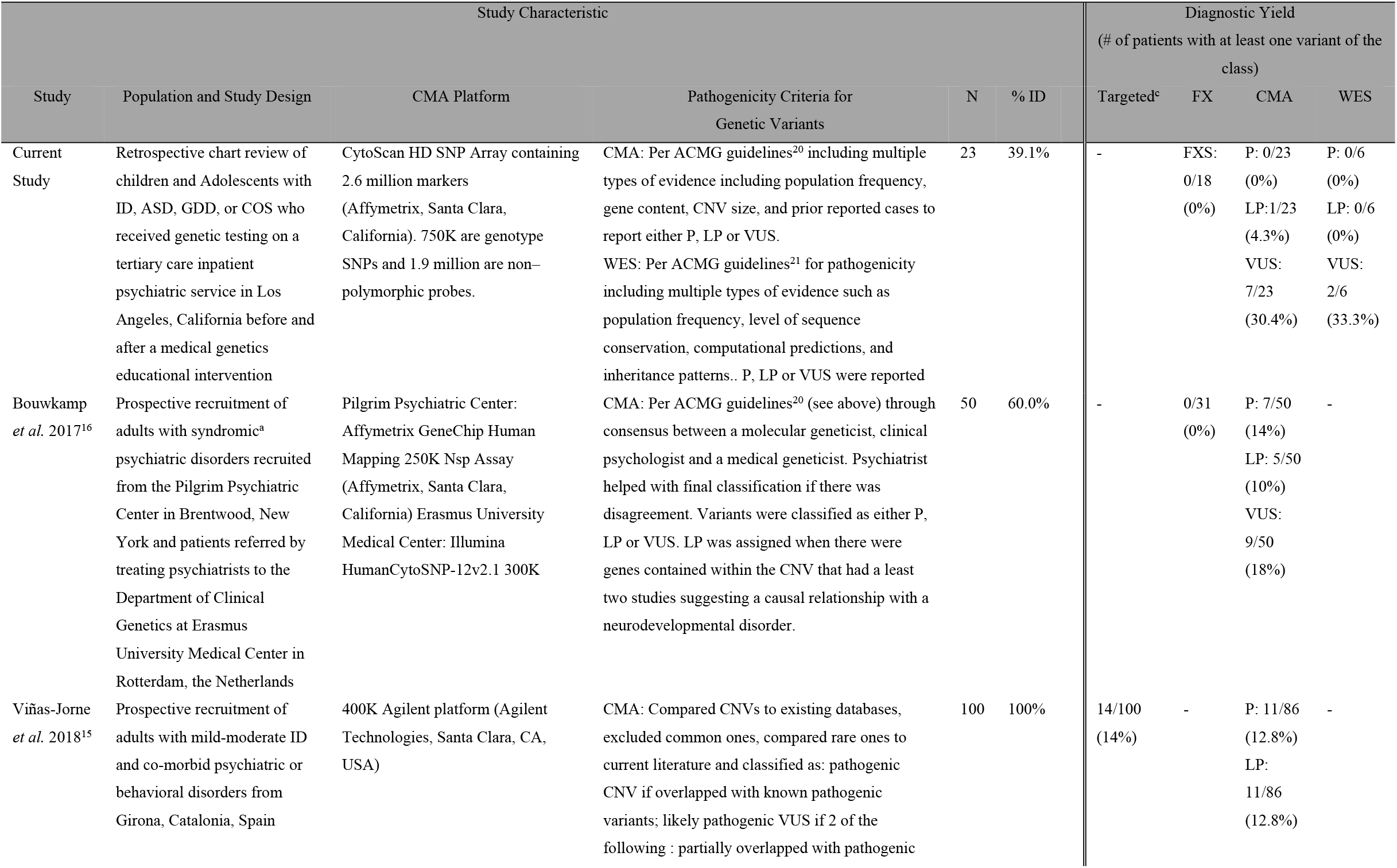
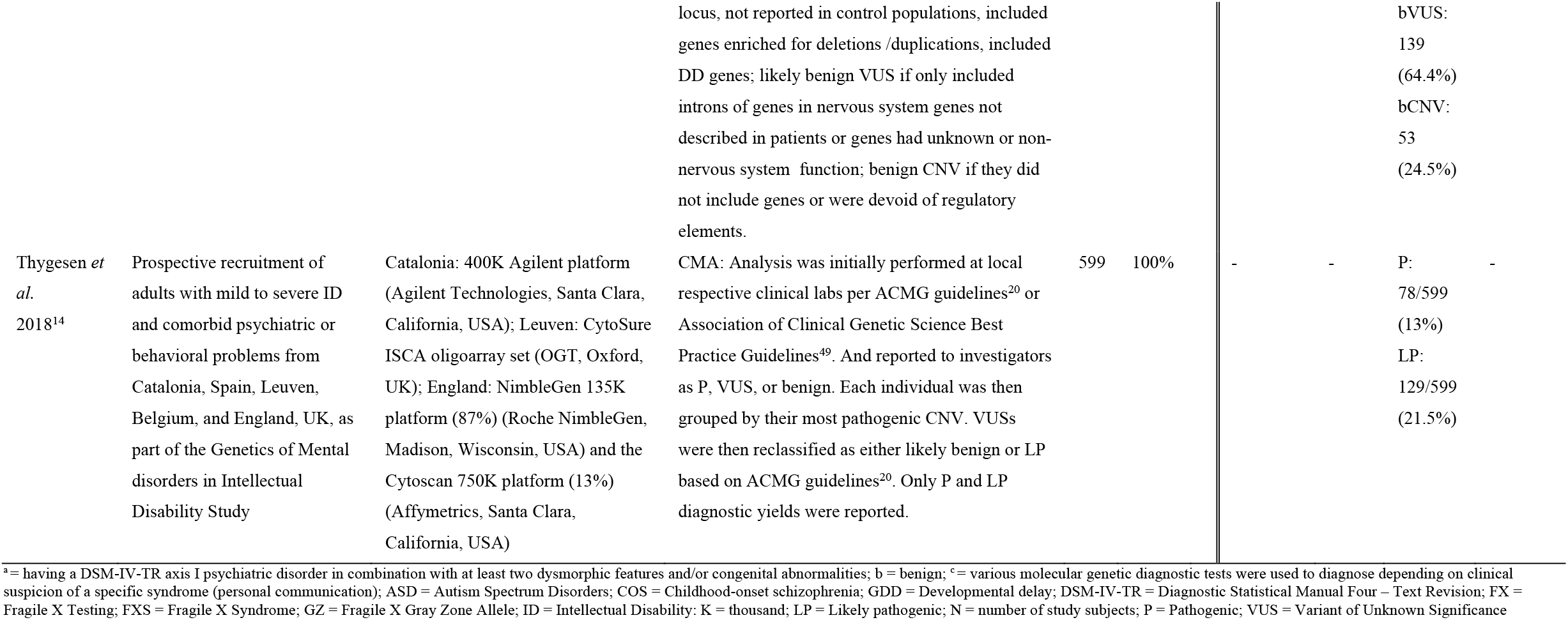
Diagnostic Yield of Genetic Testing in Patients with NDDs and Comorbid Psychiatric Disorders

The diagnostic yield of 4.3% for patients who received genetic testing while on inpatient was lower than we expected. We hypothesized a diagnostic yield of between 15%-20% based on previous studies in NDD populations^7^. Our low yield may be an artifact of low sample size, but there are other factors to consider. First, our patient population is considerably enriched for severe comorbid psychiatric disorders, especially intermittent explosive disorder (IED) and ADHD (Figure 1), and therefore may have a different genetic architecture to the general NDD population. However, previous studies of adults with ID and comorbid psychiatric disorders^14–16^ (Table 3) report a P/LP diagnostic yield of 24-34.6% (Table 3)^14–16^. There are meaningful differences between these studies and ours that likely explains some of the difference in yield. Based on the pathogenicity classification methods used (Table 3), two studies likely classified some variants as LP that we classified as VUS^14,15^. This is reflected in our high overall yield of VUSs of 34.8% (Table 1, Tables S1, S2, available online). In fact, we had two patients with the same VUS (15q11.2 duplication, breakpoints 1-2) (Table 1, Table S1, available online). Currently, its pathogenicity is unclear because of a lack of a clear association with a specific NDD phenotype^33^. However, as more patients with NDDs and psychiatric comorbidities are included in large-scale genetic studies and more affected patients with these rare variants are identified, more variants may reach the threshold for pathogenicity and the diagnostic yields between studies may start to align.

Other important difference from previous studies are the demographics and the prevalence of specific psychiatric disorders in each cohort. The previous studies had a larger percentage of patients with ID (39.1% current study vs. 60-100%^14–16^), which is a population that may be enriched for rare, pathogenic variants compared to those with ASD or GDD. The other studies also did not report the ethnic or racial breakdown of their cohorts. We had a relatively high percentage of underrepresented minorities 9/23 (39.1%) who are not well represented in most NDD genetics studies. Therefore, it is possible that these patients have less well-established disease-associated variants. Furthermore, the previous studies were in adult populations whose psychiatric and behavioral problems are somewhat different to those seen in our young population. For example, they identified a relatively high rate of genetic syndromes known to be associated with psychosis, such as 22q11.2 deletion syndrome and 16p11.2 duplication syndrome (Table 3)^14–16^. In contrast, our patients most commonly presented to the hospital due to aggressive behavior related to impulse control disorders such as ADHD and IED (Figure 1). Lastly, we reviewed the charts of all eligible patients to verify that our low yield was not due to the exclusion of patients with a pre-existing genetic diagnosis. We only identified a single patient with a pre-existing diagnosis of Klinefelter Syndrome, suggesting that this was not a major confounder. It is unclear why so few patients with genetic syndromes commonly associated with NDDs and severe behavioral problems, such as Prader-Willi Syndrome, were not hospitalized at our institution during the 31-month study period.

Lastly, inherited common variants (e.g. single nucleotide polymorphisms that are passed down from parents to child) also contribute to the genetic liability of NDDs^34–36^, although to less of a degree than in common psychiatric disorders such as OCD^37,38^, schizophrenia^39^, and depression^40^. It may be that patients with NDDs and severe psychiatric comorbidities have a genetic liability architecture more similar to common psychiatric disorders and therefore a lower burden of rare, *de novo*, pathogenic variants that we would have detected with our diagnostic approach. The high burden of mental illness in our patient’s families (Table S4, available online) supports this possibility.

### Impact on Clinical Care

One unexpected finding was that 9/23 (39.1%) of the patients tested on the inpatient service over the entire 31-month study were underrepresented minorities (Table 1), compared to 1/13 (7.7%) of those tested in other clinical settings (Table 2), with a trend towards significance (*p* = 0.057) limited by our sample size and power. This suggests that genetic testing on the inpatient CAP service may help improve access to genetic services for underrepresented minorities with NDDs^41–43^. We think this may be the case because once a child is hospitalized, several barriers to care, including insurance authorization and language and financial hurdles limiting specialty referrals, are reduced.

There were no instances where the genetic test results had a direct effect on medical management, although this has been demonstrated elsewhere^10–12,44^. The low diagnostic yield and high VUS rate limited the likelihood of the genetic testing having a direct effect on management. However, as more VUSs become reclassified as pathogenic and each disorder becomes better characterized, the chance of improving clinical care will increase. Our patient with psychosis, a unique facial gestalt, and a marfanoid habitus consistent with 3q27.2 deletion syndrome but without any pathogenic variants in that region was referred to a research study with the hopes of identifying a novel gene or genomic region associated with her unique phenotype. If it is successful, it may have implications for that patient and others in the future.

### Trainee Education and Genetic Testing Implementation

A major hurdle to the implementation of genetic testing in the inpatient CAP setting is the lack of training that many psychiatrists and psychiatry trainees have in medical genetics^13^. CAP fellow education and attending physician-driven consultation to genetic psychiatry service resulted in a significant increase in testing rate, from 1.6% to 10.7% of eligible patients (*p* = 0.0015), primarily driven by an increase in CAP fellow testing, as they tested fourteen out of the 21 patients in the post-intervention period. This jump in testing rates is consistent with our experience educating the CAP fellows on medical genetics while on their inpatient rotation. They were extremely receptive to learning about genetic testing and after every didactic session, several would approach our team with questions about how to proceed with testing for current patients of theirs. They were very proactive in reaching out to our team for additional advice on new, eligible patients. Most CAP fellows preferred that our team lead the consent process, but they were always in attendance. This level of engagement was significantly greater compared to our prior experiences giving lectures as part of standard didactic training only.

### Limitations and Future Directions

A major limitation to our study was the small number of tested patients, which reduces the generalizability of our findings. Ongoing, large-scale efforts such as those by the Autism Inpatient Collection will be essential to improve our understanding of the genetic architecture of NDDs with comorbid psychiatric disorders^45,46^. With this additional information, annual reanalysis of the genetic test results will improve diagnostic yield, as has been demonstrated in other studies of NDDs^47^. We were also limited by our lack of access to all health records outside of UCLA, thus we may have underestimated the rate of outpatient genetic testing for our patients. We attempted to address this question by searching all EHR diagnostic codes for specific genetic syndromes to capture all genetic diagnoses that were manually input by clinicians. We only identified a single patient with a known genetic syndrome (XXY, Klinefelter Syndrome) that had been diagnosed outside of our institution prior to admission. This suggests that we likely identified almost all patients with a known genetic syndrome and if patients received genetic testing elsewhere, it had likely come back negative or with VUSs that were not reported by families or not entered into the EHR by clinicians, consistent with our other findings.

Another challenge we encountered was to provide proper outpatient genetics follow-up. Outpatient follow-up is especially important for inpatient genetic testing because the majority of the time the test results come back after the patient is discharged. Explicit directions were provided in our protocol for arranging outpatient follow-up and social workers on the inpatient teams were available to assist with this process. Despite these efforts, over 40% of our patients were lost to follow-up (Figure 2), although we can’t be certain that families didn’t find genetics follow-up on their own. There were likely many contributing factors, but the primary ones we identified were challenges with insurance authorization for follow-up appointments (especially with health maintenance organizations), psychiatry trainees’ lack of familiarity with outpatient genetics services, and confusion about responsibility for follow-up. Our loss to follow-up rate may even underestimate what others might experience because two of us provide outpatient genetics follow-up services, as a medical geneticist (J.A.M.A.) and genetic psychiatrist (A.D.B.). One way to address this challenge in the future may be to provide more rapid genetic testing on the inpatient service, as it has been successfully implemented in intensive care settings^48^. If the genetic test results were available within several days (instead of weeks), they could be returned to the family prior to discharge, and potentially impact inpatient management^10–12^.

## Data Availability

All relevant data is included in the manuscript and supplement submitted.

## Acknowledgements and Funding

We would like to thank Carina Hampp, Douglas Bell, MD, PhD and the UCLA Clinical Translational Sciences Institute (CTSI) Informatics Program (funded by National Institutes of Health National Center for Advancing Translational Science CTSI Grant UL1TR001881) for all of their assistance with data extraction and preparation. We would like to thank Dr. Myung-Shin Sim from the UCLA CTSI and Department of Medicine biostatistical consultation service for her assistance with data analysis. The authors would like to thank the Genome Aggregation Database (gnomAD) and the groups that provided exome and genome variant data to this resource. A full list of contributing groups can be found at https://gnomad.broadinstitute.org/about. A.D.B. received funding support from the National Institutes of Health T32 UCLA Intercampus Medical Genetics Training Program (2T32GM008243-31). J.S. received funding support from the UCLA Medical Student Summer Research Fellowship Program in Psychiatry and Biobehavioral Sciences.

## References

1. Vorstman JAS, Parr JR, Moreno-De-Luca D, Anney RJL, Nurnberger Jr JI, Hallmayer JF. Autism genetics: opportunities and challenges for clinical translation. Nat Rev Genet. 2017;18(6):362–376. doi:10.1038/nrg.2017.4

2. Study DDD, McRae JF, Clayton S, et al. Prevalence and architecture of de novo mutations in developmental disorders. Nature. 2017;542(7642):433–438. doi:10.1038/nature21062

3. Sanders SJ, He X, Willsey AJ, et al. Insights into Autism Spectrum Disorder Genomic Architecture and Biology from 71 Risk Loci. Neuron. 2015;87(6):1215–1233. doi:10.1016/J.NEURON.2015.09.016

4. Sestan N, State MW. Lost in Translation: Traversing the Complex Path from Genomics to Therapeutics in Autism Spectrum Disorder. Neuron. 2018;100(2):406–423. doi:10.1016/J.NEURON.2018.10.015

5. Miller DT, Adam MP, Aradhya S, et al. Consensus statement: chromosomal microarray is a first-tier clinical diagnostic test for individuals with developmental disabilities or congenital anomalies. Am J Hum Genet. 2010;86(5):749–764. doi:10.1016/j.ajhg.2010.04.006

6. Muhle RA, Reed HE, Vo LC, et al. Clinical Diagnostic Genetic Testing for Individuals With Developmental Disorders. J Am Acad Child Adolesc Psychiatry. 2017;56(11):910–913. doi:10.1016/j.jaac.2017.09.418

7. Srivastava S, Love-Nichols JA, Dies KA, et al. Meta-analysis and multidisciplinary consensus statement: exome sequencing is a first-tier clinical diagnostic test for individuals with neurodevelopmental disorders. Genet Med. June 2019. doi:10.1038/s41436-019-0554-6

8. Riggs ER, Wain KE, Riethmaier D, et al. Chromosomal microarray impacts clinical management. Clin Genet. 2014;85(2):147–153. doi:10.1111/cge.12107

9. Reiff M, Giarelli E, Bernhardt BA, et al. Parents’ Perceptions of the Usefulness of Chromosomal Microarray Analysis for Children with Autism Spectrum Disorders. J Autism Dev Disord. 2015;45(10):3262–3275. doi:10.1007/s10803-015-2489-3

10. Fung WLA, Butcher NJ, Costain G, et al. Practical guidelines for managing adults with 22q11.2 deletion syndrome. Genet Med. 2015;17(8):599–609. doi:10.1038/gim.2014.175

11. Serret S, Thümmler S, Dor E, Vesperini S, Santos A, Askenazy F. Lithium as a rescue therapy for regression and catatonia features in two SHANK3 patients with autism spectrum disorder: case reports. BMC Psychiatry. 2015;15(1):107. doi:10.1186/s12888-015-0490-1

12. De Leersnyder H, de Blois MC, Vekemans M, et al. beta(1)-adrenergic antagonists improve sleep and behavioural disturbances in a circadian disorder, Smith-Magenis syndrome. J Med Genet. 2001;38(9):586–590. doi:10.1136/jmg.38.9.586

13. Besterman AD, Moreno-De-Luca D, Nurnberger JI. 21st-Century Genetics in Psychiatric Residency Training. JAMA Psychiatry. 2019;76(3):231. doi:10.1001/jamapsychiatry.2018.3872

14. Thygesen JH, Wolfe K, McQuillin A, et al. Neurodevelopmental risk copy number variants in adults with intellectual disabilities and comorbid psychiatric disorders. Br J Psychiatry. 2018;212(5):287–294. doi:10.1192/bjp.2017.65

15. Viñas-Jornet M, Esteba-Castillo S, Baena N, et al. High Incidence of Copy Number Variants in Adults with Intellectual Disability and Co-morbid Psychiatric Disorders. Behav Genet. 2018;48(4):323–336. doi:10.1007/s10519-018-9902-6

16. Bouwkamp CG, Kievit AJA, Markx S, et al. Copy Number Variation in Syndromic Forms of Psychiatric Illness: The Emerging Value of Clinical Genetic Testing in Psychiatry. Am J Psychiatry. 2017;174(11):1036–1050. doi:10.1176/appi.ajp.2017.16080946

17. Kalia SS, Adelman K, Bale SJ, et al. Recommendations for reporting of secondary findings in clinical exome and genome sequencing, 2016 update (ACMG SF v2.0): a policy statement of the American College of Medical Genetics and Genomics. Genet Med. 2017;19(2):249–255. doi:10.1038/gim.2016.190

18. Lee H, Deignan JL, Dorrani N, et al. Clinical Exome Sequencing for Genetic Identification of Rare Mendelian Disorders. JAMA. 2014;312(18):1880. doi:10.1001/jama.2014.14604

19. Lek M, Karczewski KJ, Minikel E V., et al. Analysis of protein-coding genetic variation in 60,706 humans. Nature. 2016;536(7616):285–291. doi:10.1038/nature19057

20. Kearney HM, Thorland EC, Brown KK, Quintero-Rivera F, South ST, Working Group of the American College of Medical Genetics Laboratory Quality Assurance Committee. American College of Medical Genetics standards and guidelines for interpretation and reporting of postnatal constitutional copy number variants. Genet Med. 2011;13(7):680–685. doi:10.1097/GIM.0b013e3182217a3a

21. Richards S, Aziz N, Bale S, et al. Standards and guidelines for the interpretation of sequence variants: a joint consensus recommendation of the American College of Medical Genetics and Genomics and the Association for Molecular Pathology. 2015. doi:10.1038/gim.2015.30

22. Campbell I. Chi-squared and Fisher–Irwin tests of two-by-two tables with small sample recommendations. Stat Med. 2007;26(19):3661–3675. doi:10.1002/sim.2832

23. Ahn K, Gotay N, Andersen TM, et al. High rate of disease-related copy number variations in childhood onset schizophrenia. Mol Psychiatry. 2014;19(5):568–572. doi:10.1038/mp.2013.59

24. Ambalavanan A, Girard SL, Ahn K, et al. De novo variants in sporadic cases of childhood onset schizophrenia. Eur J Hum Genet. 2016;24(6):944–948. doi:10.1038/ejhg.2015.218

25. Ambalavanan A, Chaumette B, Zhou S, et al. Exome sequencing of sporadic childhood-onset schizophrenia suggests the contribution of X-linked genes in males. Am J Med Genet Part B Neuropsychiatr Genet. October 2018. doi:10.1002/ajmg.b.32683.

26. Chaumette B, Ferrafiat V, Ambalavanan A, et al. Missense variants in ATP1A3 and FXYD gene family are associated with childhood-onset schizophrenia. Mol Psychiatry. June 2018:1. doi:10.1038/s41380-018-0103-8

27. Peykov S, Berkel S, Schoen M, et al. Identification and functional characterization of rare SHANK2 variants in schizophrenia. Mol Psychiatry. 2015;20(12):1489–1498. doi:10.1038/mp.2014.172

28. Homann OR, Misura K, Lamas E, et al. Whole-genome sequencing in multiplex families with psychoses reveals mutations in the SHANK2 and SMARCA1 genes segregating with illness. Mol Psychiatry. 2016;21(12):1690–1695. doi:10.1038/mp.2016.24

29. Howrigan DP, Rose SA, Samocha KE, et al. Schizophrenia risk conferred by protein-coding de novo mutations. bioRxiv. December 2018:495036. doi:10.1101/495036

30. Thevenon J, Callier P, Poquet H, et al. 3q27.3 microdeletional syndrome: a recognisable clinical entity associating dysmorphic features, marfanoid habitus, intellectual disability and psychosis with mood disorder. J Med Genet. 2014;51(1):21–27. doi:10.1136/jmedgenet-2013-101939

31. Fukushi D, Yamada K, Suzuki K, et al. Clinical and genetic characterization of a patient with SOX5 haploinsufficiency caused by a de novo balanced reciprocal translocation. Gene. 2018;655:65–70. doi:10.1016/J.GENE.2018.02.049

32. Mullegama S V, Klein SD, Nguyen DC, et al. Is it time to retire fragile X testing as a first-tier test for developmental delay, intellectual disability, and autism spectrum disorder? Genet Med. 2017;19(12):1380–1380. doi:10.1038/gim.2017.146

33. Mohan KN, Cao Y, Pham J, et al. Phenotypic association of 15q11.2 CNVs of the region of breakpoints 1–2 (BP1–BP2) in a large cohort of samples referred for genetic diagnosis. J Hum Genet. 2019;64(3):253–255. doi:10.1038/s10038-018-0543-7

34. Niemi MEK, Martin HC, Rice DL, et al. Common genetic variants contribute to risk of rare severe neurodevelopmental disorders. Nature. 2018;562(7726):268–271. doi:10.1038/s41586-018-0566-4

35. Gaugler T, Klei L, Sanders SJ, et al. Most genetic risk for autism resides with common variation. Nat Genet. 2014;46(8):881–885. doi:10.1038/ng.3039

36. Weiner DJ, Wigdor EM, Ripke S, et al. Polygenic transmission disequilibrium confirms that common and rare variation act additively to create risk for autism spectrum disorders. Nat Genet. 2017;49(7):978–985. doi:10.1038/ng.3863

37. Mattheisen M, Samuels JF, Wang Y, et al. Genome-wide association study in obsessive-compulsive disorder: results from the OCGAS. Mol Psychiatry. 2015;20(3):337–344. doi:10.1038/mp.2014.43

38. (OCGAS) IOCDFGC (IOCDF-G and OCGAS, Arnold PD, Askland KD, et al. Revealing the complex genetic architecture of obsessive–compulsive disorder using meta-analysis. Mol Psychiatry. 2018;23(5):1181–1188. doi:10.1038/mp.2017.154

39. Schizophrenia Working Group of the Psychiatric Genomics Consortium SWG of the PG, Ripke S, Neale BM, et al. Biological insights from 108 schizophrenia-associated genetic loci. Nature. 2014;511(7510):421–427. doi:10.1038/nature13595

40. Wray NR, Ripke S, Mattheisen M, et al. Genome-wide association analyses identify 44 risk variants and refine the genetic architecture of major depression. Nat Genet. 2018;50(5):668–681. doi:10.1038/s41588-018-0090-3

41. Shea L, Newschaffer CJ, Xie M, Myers SM, Mandell DS. Genetic testing and genetic counseling among medicaid-enrolled children with autism spectrum disorder in 2001 and 2007. Hum Genet. 2014;133(1):111–116. doi:10.1007/s00439-013-1362-8

42. Cuccaro ML, Czape K, Alessandri M, et al. Genetic testing and corresponding services among individuals with autism spectrum disorder (ASD). Am J Med Genet Part A. 2014;164(10):2592–2600. doi:10.1002/ajmg.a.36698

43. Smith AJ, Oswald D, Bodurtha J. Trends in Unmet Need for Genetic Counseling Among Children With Special Health Care Needs, 2001–2010. Acad Pediatr. 2015;15(5):544–550. doi:10.1016/J.ACAP.2015.05.007

44. Coulter ME, Miller DT, Harris DJ, et al. Chromosomal microarray testing influences medical management. Genet Med. 2011;13(9):770–776. doi:10.1097/GIM.0b013e31821dd54a

45. Siegel M. The Severe End of the Spectrum: Insights and Opportunities from the Autism Inpatient Collection (AIC). J Autism Dev Disord. 2018;48(11):3641–3646. doi:10.1007/s10803-018-3731-6

46. Siegel M, Smith KA, Mazefsky C, et al. The autism inpatient collection: methods and preliminary sample description. Mol Autism. 2015;6(1):61. doi:10.1186/s13229-015-0054-8

47. Wright CF, McRae JF, Clayton S, et al. Making new genetic diagnoses with old data: iterative reanalysis and reporting from genome-wide data in 1,133 families with developmental disorders. Genet Med. 2018;20(10):1216–1223. doi:10.1038/gim.2017.246

48. Saunders CJ, Miller NA, Soden SE, et al. Rapid Whole-Genome Sequencing for Genetic Disease Diagnosis in Neonatal Intensive Care Units. Sci Transl Med. 2012;4(154):154ra135–154ra135. doi:10.1126/scitranslmed.3004041

49. Ellard S, Baple EL, Owens M, et al. ACGS Best Practice Guidelines for Variant Classification 2017. https://www.acgs.uk.com/media/10792/uk_practice_guidelines_for_variant_classification_2017.pdf. Published 2017.

